# Multimodal sleep stage classification and label-free abnormality scoring in mid-to-older adults

**DOI:** 10.64898/2026.05.28.26353980

**Authors:** Zekeriye Nur, Nivedita Bijlani, Mauricio Villarroel

## Abstract

**Background:** Sleep fragmentation and reduced sleep efficiency are markers of disrupted sleep architecture linked to cognitive and age-related decline. Current assessments rely on subjective reports prone to recall bias, limiting their effectiveness for longitudinal monitoring. Data-driven analysis of sleep using physiological signals such as EEG and EMG remains underutilised, particularly in mid-to-older adults.

**Objective:** We present a deep learning pipeline for automated sleep staging and label-free abnormality scoring, with the primary objective of quantifying deviations in sleep architecture to capture progressive sleep disruption and longitudinal change.

**Methods:** Temporal and attention-based models were benchmarked using datasets from the National Sleep Research Resource and PhysioBank. To improve class-specific performance, we introduce a stacking-based ensemble of sleep stage classifiers, each trained to specialise in a different stage. For longitudinal scoring, we develop a reconstruction loss-based abnormality metric using a temporal convolutional autoencoder trained on hypnograms generated by the sleep staging models.

**Results:** Attention-based models, particularly AttnSleep, achieved the highest performance in both multimodal and single-channel settings (accuracy: 0.85 and 0.83; F1: 0.79 and 0.74, respectively). The encoder-decoder ensemble model improved overall classification accuracy by 3% compared to the best-performing biased base classifier, with a modest gain in N1-stage F1 score (0.444). The proposed abnormality score correlated with Pittsburgh Sleep Quality Index components and showed sensitivity to synthetic hypnogram degradation, highlighting its potential as a label-free indicator of sleep disruption.

**Conclusion:** Automated classification and annotation-free scoring enable an end-to-end multimodal pipeline that supports scalable, objective sleep health monitoring, with relevance for future clinical deployment.

**Graphical Abstract:** 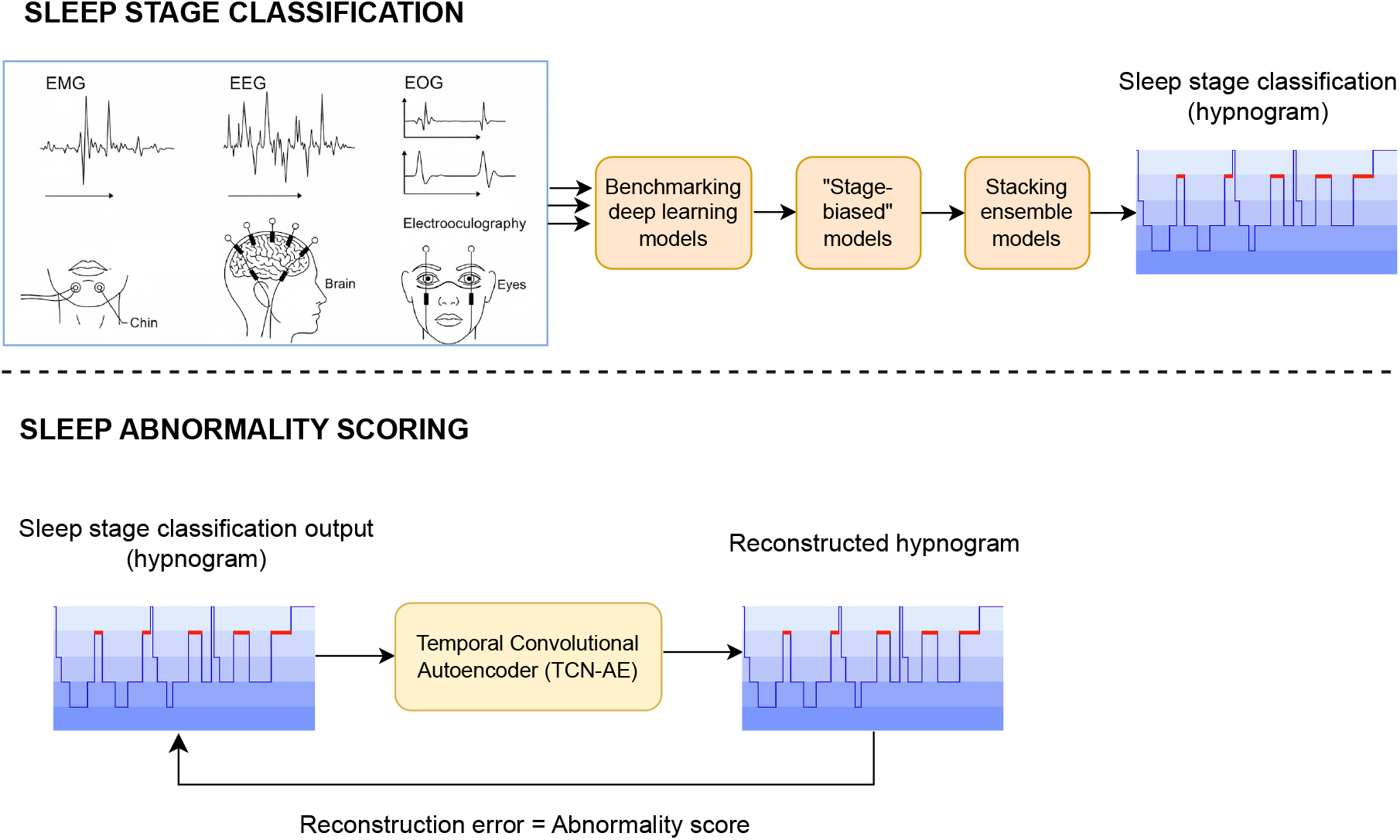

**Highlights:**

- A label-free framework quantifies sleep abnormality from hypnogram structure.
- Attention-based models outperform CNNs for sleep classification in mid-to-older adults.
- A stacking-based ensemble of stage-biased models improves N1 classification.
- Sleep abnormality scores derived from hypnograms align with PSQI indices.

## 1. Introduction

Sleep has numerous health benefits. In slow-wave sleep (SWS), growth hormones are secreted to support muscle repair, tissue growth, and cellular regeneration [1]. The glymphatic system clears toxins like beta-amyloid from the brain, with build-up associated with cognitive decline [2]. Sleep also supports memory consolidation by strengthening neural connections and reorganising synaptic pathways [3] [4]. Disrupted sleep architecture and abnormal sleep stage transitions are often associated with cognitive and age-related decline.

Sleep comprises two main stages: Non-rapid eye movement (NREM) and rapid eye movement (REM), each supporting distinct restorative functions. According to American Academy of Sleep Medicine (AASM) guidelines [5], NREM is further divided into three stages: N1 (light sleep), N2 (50% of total sleep, marked by K-complexes and spindles), and N3 (deep stage, critical for physical restoration and toxin clearance). REM sleep typically follows a cycle of non-REM stages and plays key roles in memory consolidation, emotional regulation, and other neural restorative processes [6] [7]. Early in the night, NREM dominates; later, REM sleep becomes more prominent. Hypnograms, which capture sleep architecture, follow generic, repeating patterns. Deviations from these patterns may indicate abnormality.

Disruptions to sleep architecture, such as increased sleep fragmentation, reduced slow-wave sleep, and altered stage transitions, have been associated with cognitive decline and adverse ageing outcomes in ageing and clinical populations [8] [9] [10]. These changes highlight the importance of objective, quantitative analysis of sleep structure for identifying and monitoring deviations from normative patterns over time.

Video-Polysomnography (vPSG) is the gold standard for sleep assessment, using multiple sensors and manual scoring based on AASM rules. Signals include video, electroencephalography (EEG), electromyography (EMG), electrooculography (EOG), electrocardiography (ECG), and pulse oximetry. Standard sleep staging is conventionally performed on 30-second epochs of physiological data, as defined by AASM scoring rules. However, vPSG is expensive, intrusive, and labour-intensive, typically requiring overnight stay in a sleep clinic and sleep specialists for scoring. Manual annotation not only introduces inter- and intra-scorer variability but also prevents fully automated, label-free analysis. Moreover, the sleep-lab setting may distort natural sleep, and a single in-lab night provides only a snapshot, missing long-term trends and night-to-night variability. These limitations underscore the clinical need for scalable, home-based systems that can integrate multimodal physiological and behavioral signals to support objective, continuous, and expert-free sleep staging and long-term monitoring.

Over the past two decades, automatic sleep classification has advanced significantly, with growing interest in multimodal models that integrate EEG, ECG, and EMG signals. A recent study [11] tackles the clinical need for automated, label-free sleep staging and disorder screening. The authors propose a Multimodal and Multilabel Decision-Making System (MML-DMS) designed to jointly classify six sleep stages (W, N1–3, REM1-2) and eight sleep disorders including bruxism, insomnia, narcolepsy, REM behaviour disorder, and sleep-disordered breathing from the PhysioNet CAP database of 108 subjects. The pipeline converts 10 s overlapping blocks of synchronised EEG, ECG, and EMG into logarithmic Short-Time Fourier Transform (STFT) spectrograms, then RGB images. Six VGG16 convolutional neural networks (CNNs) – three for sleep staging and three for disorder detection – act as first-level single-modality classifiers. Their probability vectors are concatenated and fed into two shallow decision networks that fuse both multimodal and multilabel information to produce final stage and disorder outputs. The full MML-DMS2 configuration achieved 94.3% accuracy (F1=0.92) for sleep stage classification and 99.1% accuracy (F1=0.99) for disorder detection, outperforming single-modality and earlier multimodal methods. While methodologically strong, the approach has limitations. All data comes from a single curated dataset with overlapping subject representation between training and testing, raising concerns of data leakage and limited generalisability to new patients, home environments, or wearable devices. The spectrogram-to-image preprocessing captures time–frequency characteristics but increases computational load and may limit real-time deployment. Moreover, model training is supervised and requires large, well-labelled datasets, which conflicts with the long-term goal of fully label-free sleep staging.

Ellis et al. [12] addressed the need for interpretable multimodal sleep staging, where most deep learning methods act as black boxes and standard ablation approaches create unrealistic samples. They trained a 1D CNN on EEG, EOG, and EMG from the PhysioNet Sleep-EDF database (44 nights, 22 subjects, 42,218 thirty-second epochs) to classify five sleep stages (Awake, N1–3, REM). To improve explainability, they introduced a domain-specific ablation method that replaces each modality with synthetic line-noise (40 Hz sinusoid plus Gaussian noise) instead of simply zeroing out signals, thereby better reflecting realistic electrophysiological artefacts. This method produced importance rankings consistent with AASM scoring guidelines, while modestly accentuating the contribution of some modalities compared with zero-out ablation. Despite this innovation, the CNN achieved only moderate performance (e.g. lowest F1 on N1), and evaluation was limited to 17 training subjects with 10-fold cross-validation, raising concerns over generalisability.

Lee et al. [13] introduced *SleepXViT*, an explainable ViT (Vision Transformer) framework for automatic sleep staging using full multimodal PSG data. Unlike prior models limited to single-channel EEG or coarse CNN-based visual features, *SleepXViT* converts synchronised PSG signals (EEG, EOG, EMG, ECG, and respiratory channels) into standardised 30 s images and applies a two-stage ViT architecture. An intra-epoch ViT extracts features and classifies each epoch, while an inter-epoch ViT captures temporal dependencies across neighbouring epochs. The model achieved strong performance on a large, diverse KISS dataset (7,745 overnight studies) with a macro F1 of 81.9%, and remained performant on external SHHS datasets. Explainability is provided through well-calibrated confidence scores and high-resolution relevance heatmaps that align with AASM staging criteria and quantify the influence of adjacent epochs. Despite these strengths, the approach relies on spectrogram-to-image preprocessing, which introduces latency and computational overhead, limiting its suitability for real-time or low-power wearable deployment.

In several works, age is a critical and often-overlooked variable. A sleep pattern typical for older adults may be atypical in younger individuals; thus, excluding age from classification models introduces potential misclassifications. In longitudinal monitoring, this age-influenced drift may obscure clinically relevant deviations, especially in ageing populations. While extensive research exists on sleep stage detection, annotation-free sleep abnormality quantification using multimodal data remains unexplored. Current subjective approaches for sleep assessment, such as the Pittsburgh Sleep Quality Index (PSQI) [14], are inherently subject to recall bias.

This study addresses key gaps in multimodal sleep analysis for mid-to-older adults, an age group exhibiting substantial variability in sleep architecture and longitudinal change. The primary aim of this work is to develop a label-free method for quantifying sleep abnormality from hypnogram structure, with sleep stage classification serving as an enabling component within an end-to-end pipeline. Our proposed method establishes foundational components for sleep abnormality quantification from raw physiological signals. We develop an autoencoder-based sleep abnormality scoring method that operates directly on a subject’s hypnogram, without requiring prior labelling by sleep experts, providing an objective and repeatable approach for longitudinal sleep monitoring. Sleep abnormality is quantified from the reconstruction loss of the hypnogram, with the autoencoder learning the temporal patterns characteristic of normative sleep and highlighting deviations from these patterns. The key contributions of this work are summarised below:

- We propose a novel, label-free sleep abnormality scoring model that operates directly on hypnograms, enabling objective sleep assessment.
- We benchmark deep learning models for multimodal sleep stage classification in mid-to-older adults using large, heterogeneous public datasets.
- We develop a stage-specialised ensemble model to improve class-specific classification performance, particularly for challenging stages such as N1.

## 2. Methods

### 2.1. Datasets

We used the following three datasets in this study:

#### Sleep Heart Health Study (SHHS) [15]

A multi-centre cohort study implemented by the National Heart, Lung, and Blood Institute to investigate the cardiovascular and other consequences of sleep-disordered breathing. It contains data from 6441 adults aged 40 years and older. Data were collected over two exam cycles, with cardiovascular outcomes tracked until 2010. The following signals were acquired from the recording montage: C3/A2 and C4/A1 EEGs sampled at 125 Hz, right and left EOGs sampled at 50 Hz, and a bipolar submental (chin) EMG sampled at 125 Hz.

#### Wisconsin Sleep Cohort (WSC) [16]

An ongoing longitudinal study examining the causes, consequences and natural history of sleep disorders, particularly sleep apnoea. It uses overnight in-laboratory sleep studies conducted with a baseline sample of 1500 Wisconsin state employees, assessed at four-year intervals. This study uses O1-M2 and C3-M2 EEG channels, left and right EOGs, and bipolar submental (chin) EMG sampled at 100 Hz.

#### Sleep-EDF-20 and Sleep-EDF-78 [17]

Whole-night polysmnographic sleep recordings from 153 Caucasian individuals aged 25–101 years. The corresponding hypnograms contain annotations of the sleep stages that correspond to the PSGs. All hypnograms were manually scored by well-trained technicians. This study uses Fpz-Cz and Pz-Oz EEG, horizontal EOG, and submental chin EMG signals sampled at 100 Hz.

Each dataset was evaluated independently, and performance metrics were averaged across datasets. Within each dataset, we also examined differences between single-channel EEG input and multimodal inputs (EEG, EOG, and EMG). The EOG and EMG channels are consistent across datasets, whereas the EEG channels differ. Specifically, SHHS includes C3/A2 and C4/A1, Sleep-EDF includes Fpz-Cz and Pz-Oz, and WSC includes O1-M2 and C3-M2. Despite these differences, all selected channels correspond to standard scalp locations (frontal, central, or occipital) used in sleep staging and capture key electrophysiological features such as sleep spindles and slow-wave activity.

### 2.2. Recording selection

We filtered recordings to focus on individuals aged 40+ years. Included recordings contained ≥ 5% N3 sleep, ≥ 15% REM sleep, and a total sleep time of ≥ 6 hours. We excluded recordings with substantial signal artefacts and individuals with diagnosed neurological or psychiatric sleep disorders. For datasets containing subjects with sleep-disordered breathing (e.g., SHHS and WSC), the Apnoea–Hypopnoea Index (AHI) was used as a standard clinical measure of sleep pathology, with AHI *<* 5 events/hour considered indicative of normal sleep.

Epochs marked “unknown” or “movement” were removed by the clinicians. N3 and N4 sleep stages were merged into a single N3 stage, consistent with AASM guidelines to simplify scoring and enhance inter-rater reliability for deep slow-wave sleep. Wake periods before sleep onset and after final awakening were truncated to 30 minutes to mitigate class imbalance.

For each sleep stage (excluding wake), the standard deviation of each epoch was calculated. Artefacts were defined as epochs with values exceeding 3 standard deviations from the stage-wise mean. When multiple channels were used, artefact detection was performed via majority voting across channels. Recordings with more than 2% artefactual epochs were excluded, as a small proportion of artefacts is common in PSG data, while higher proportions can distort sleep stage distributions and bias model training [5] [18]. The 2% threshold was selected as a conservative quality-control criterion to retain the majority of recordings while removing those with substantial signal corruption. Epochs identified as artefactual were removed and not replaced; no synthetic reconstruction or interpolation was performed. After artefact removal, 521 recordings from 501 subjects remained, with the following class distribution: W: 26,607; N1: 11,141; N2: 63,595; N3: 15,025; REM: 22,273.

Figure 1 shows the age distribution of the combined cohort.

**Figure 1:**
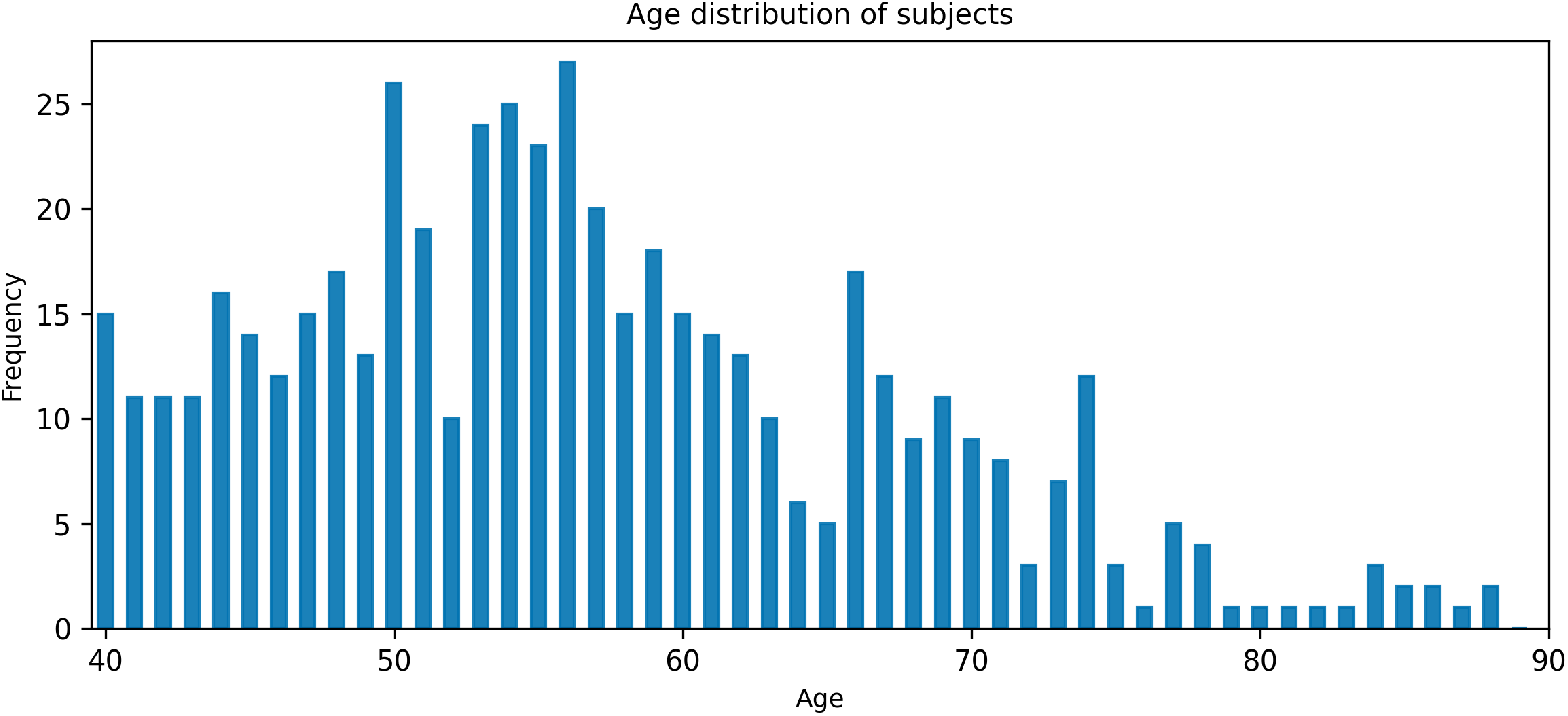
Age distribution of participants aged 40 years and above, across the SHHS, WSC, and Sleep-EDF datasets.

### 2.3. Preprocessing

The SHHS data, with a sample frequency of 125 Hz, was downsampled to match the 100 Hz sampling of the WSC and Sleep-EDF datasets. To mitigate the risk of aliasing, a polyphase filter was utilised to attenuate all frequencies above 50 Hz. In order to attenuate low-frequency noise due to sweating, electrode drift or movement artefacts, and high frequency noise due to muscle activity, sudden movements, eyelid fluttering or external electromagentic interference, bandpass filtering was applied to filter EEG and EOG signals between 0.3–30 Hz, and EMG signals between 10–50 Hz.

### 2.4. Sleep stage classification

A range of time series models combining CNNs, Recurrent Neural Networks (RNNs), and Transformer-based architectures were selected to capture temporal dependencies and classify sleep stages.

#### 2.4.1. Base models

To capture spatial features and temporal dynamics from physiological signals, we implemented the following deep learning architectures for sleep stage classification:

##### ChronoNet

[19] — This model combines multiple 1D convolutional layers for spatial feature extraction with deep Gated Recurrence Units (GRUs) to model temporal dependencies. Convolutional blocks use kernel sizes of 2, 4, and 8 to capture multi-scale temporal features, which are stacked and passed to downstream GRU layers. GRUs are arranged with skip connections to ensure efficient information flow across layers.

##### Hybrid-MultiChannelNet

[20]— Designed specifically for multimodal physiological time series, this model employs a lightweight, linear architecture optimised for sleep staging. The first part of the model consists of four convolutional layers to extract deep features from the multimodal data stream. The extracted features from the CNN are fed into a GRU to learn the sequential trends and long-term dependencies in the signal. The outputs from the GRU are passed to a fully connected layer. Finally, a Softmax layer is used to classify the output from the fully connected layer into five distinct sleep stages: Wake (W), Stage 1 (N1), Stage 2 (N2), Stage 3 (N3), and REM.

##### SleepContextNet

[21] — This is a sequence-to-sequence model designed for automatic sleep staging from a single-channel EEG. The model processes individual 30-second EEG epochs through a CNN to extract relevant features, including specific wave patterns and frequency components. The CNN is composed of three convolutional layers, a depthwise convolution, and two Channel Spatial Attention (1D-CSA) layers to enhance feature learning. These layers are based on the idea of Continuous Block Attention Module (CBAM) [22], an attention mechanism designed to refine feature maps in CNNs. A 1D CSA layer is a CBAM modified for one-dimensional signals, with the features adaptively adjusted in the channel and spatial dimensions, to enable the network to pay more attention to the prominent features of EEG signals. The extracted features are then passed sequentially to a single Long Short-Term Memory (LSTM) layer, responsible for learning both short-term dependencies and long-term temporal context to improve the accuracy of the final sleep stage classification.

##### ConvTransSleepNet

[23] — This model integrates a Multi-Resolution CNN (MCRNN) with Transformer encoders. The MCRNN comprises two convolutional branches with different kernel sizes – a narrow kernel to capture high-frequency features, and a wider kernel to extract low-frequency patterns. Their outputs are concatenated and passed to a Transformer block composed of six encoder layers to model long-range temporal dependencies. A fully connected layer produces the final output.

##### AttnSleep

[24] — This model combines the MCRNN backbone with Adaptive Feature Recalibration (AFR), an attention-based mechanism that dynamically adjusts the importance of feature maps via global pooling and fully connected layers. AttnSleep uses a Temporal Context Encoder (TCE) to capture sequence dependencies. The architecture includes an MCRNN, AFR module, two TCE blocks, and a fully connected output layer. AFR enables the network to prioritise the most relevant features, enhancing discriminative power for sleep stage classification.

We further investigated whether integrating an attention mechanism into the best-performing non-attention architecture could enhance its classification performance.

#### 2.4.2. Training

Model parameters were initialised using Xavier initialisation [25] for linear layers and Kaiming initialisation [26] for convolutional layers. The Adam optimiser [27] was used to adaptively learn the model parameters. As sleep datasets are typically class-imbalanced, we used a class-aware cross-entropy (CACE) loss by multiplying the loss per sample with a weight associated with the true class.

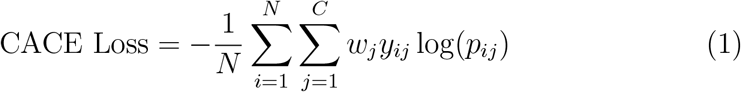

where *N* is the number of training samples, *C* is the number of classes or sleep stages, *y*_*ij*_ is the one-hot encoded ground truth label so that *y*_*ij*_ = 1 if sample *i* belongs to class *j* otherwise *y*_*ij*_ = 0, *p*_*ij*_ is the predicted probability that sample *i* belongs to class *j* computed using softmax, and *w*_*j*_ is the class weight associated with class *j* introduced to mitigate class imbalance.

We implemented the class-aware loss function from Eldele et al. [24]. This method accounts for both the relative density and the distinctiveness of each class. We trained our model for up to 200 epochs with early stopping (patience=15) and a `ReduceLROnPlateau` scheduler [28] (factor=10, patience=5).

Data were split by subject into training (70%), validation (20%), and test (10%) sets, ensuring no data leakage. The validation set was used for model selection, including early stopping and hyperparameter tuning. Five-fold cross-validation was applied within the training set to improve robustness and reduce variance in model performance, with performance metrics averaged across folds. Final model performance was evaluated on the held-out test set.

#### 2.4.3. Ensemble Models

We explored Transformer-based ensemble methods to improve the performance of the best-performing base models, with a specific focus on enhancing N1 classification. The transitional N1 sleep stage is notoriously difficult to score accurately, yet its duration is often an indicator of sleep fragmentation. Improving its detection can provide a more precise measure of sleep continuity and aid in the assessment of sleep disorders characterised by frequent arousals or disrupted sleep architecture. We selected Transformer-based stacking models for their capacity to capture complex temporal dependencies in sleep dynamics. To isolate the contribution of ensembling, both encoder-only and encoder–decoder architectures were evaluated on single-channel inputs. The stacking ensemble combines predictions from multiple base models via a meta-model trained on out-of-fold predictions [29]. To maximise classification performance across all sleep stages, we adapted the top-performing architectures from earlier experiments into class-biased base models, each specialised for a single stage [30]. These base models were trained on class-specific subsets of data to produce high-confidence predictions for their respective classes, while other models might predict the true class with lower certainty. Their outputs were stacked to form a new feature set used to train the meta-model.

Stacking promotes model diversity and allows the meta-model to correct systematic errors made by individual base models, enhancing classification robustness. The `ReduceLROnPlateau` scheduler was modified to track classwise validation accuracy rather than global loss, further reinforcing each model’s focus on its designated class. This strategy enables each base model to learn class-specific features, while the meta-model leverages prediction confidence to integrate outputs effectively, improving both accuracy and generalisation. The Transformer meta-model uses attention to identify informative base predictions and positional encoding to capture temporal structure. Two variants were developed: two attention heads and a 256-unit feed-forward network (FFN) as shown in Figure 2. The input shape is (epochs, classes *×* base models).

**Figure 2:**
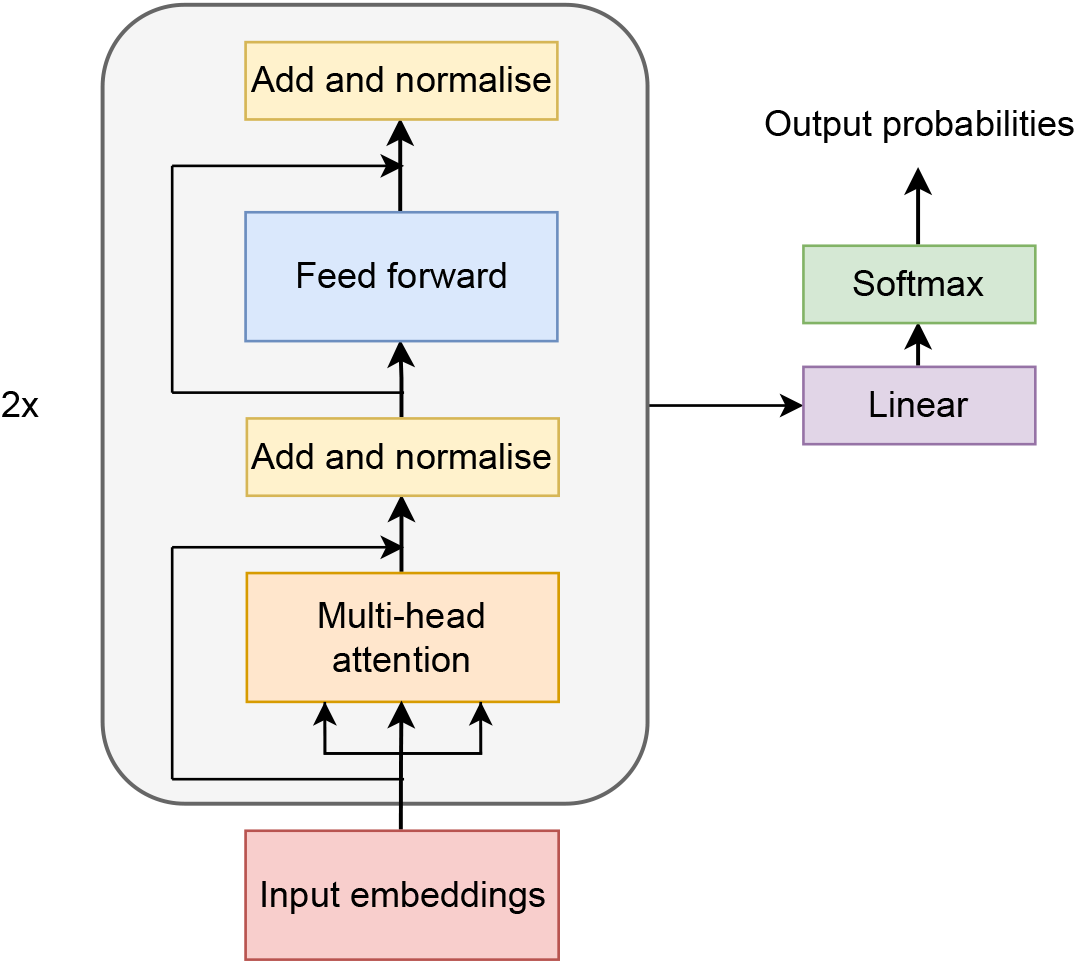
Block diagram of the encoder-only Transformer meta model based on [31].

**Encoder-only model** — This model consists of two encoder layers with

**Encoder-decoder model** — This model shares the same encoder configuration as the encoder-only model, with two additional decoder layers comprising masked self-attention, encoder-decoder attention, and FFNs, as shown in Figure 3. The input shape is: (subjects, epochs per subject, classes *×* base models). To standardise input length, we padded recordings by repeating the final epoch, under the assumption that the final 30 minutes are spent awake (Section 2.2). Positional encoding helps retain intra-subject temporal structure and align predictions to their sleep cycle. Unlike per-epoch models, this architecture operates on full-night hypnograms, enabling holistic inference. Consequently, the dataset becomes considerably smaller, as each training sample corresponds to a full-night sequence rather than a 30-second epoch.

**Figure 3:**
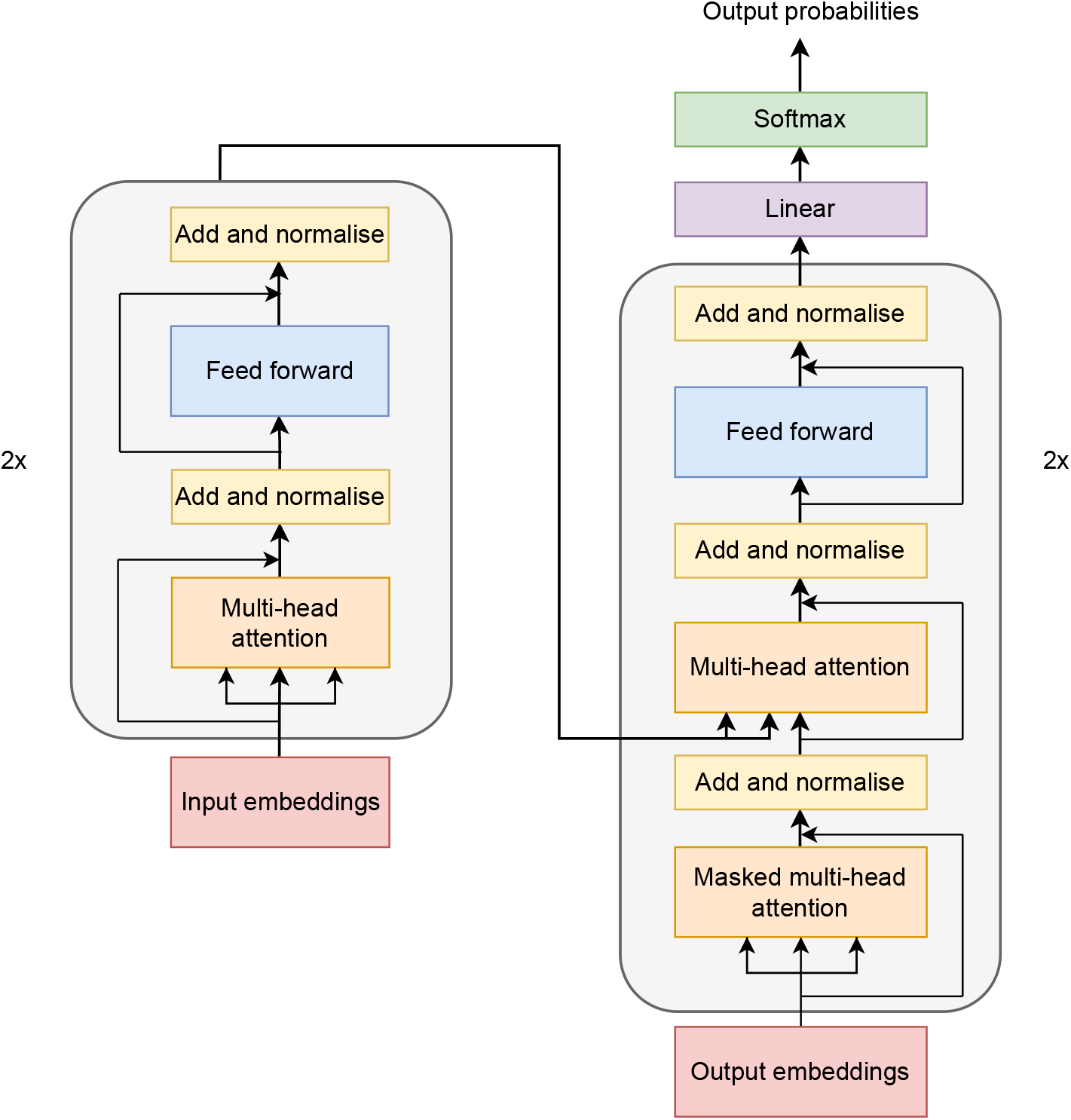
Block diagram of the encoder-decoder Transformer meta model based on [31].

### 2.5. Sleep Scoring

#### 2.5.1. Sleep abnormality score

Sleep abnormality can be understood as any significant deviation in the sequential structure, duration, or frequency of sleep stages from normative sleep architecture. The sleep abnormality score quantitatively assesses this deviation by measuring the extent to which an individual’s sleep pattern (hypnogram) differs from a model’s learned representation of healthy, standard sleep.

We developed an autoencoder-based sleep abnormality scoring method used directly on a subject’s hypnogram, without prior labelling by sleep experts. This provides an objective and repeatable scoring method for longitudinal sleep monitoring. We quantified sleep abnormality based on the reconstruction loss of the hypnogram using autoencoders, which are trained to extract salient temporal features necessary for reconstructing normative sleep stage patterns. Autoencoders represent input data into a lower-dimensional latent representation, from which the original input is reconstructed. We define the reconstruction loss as the difference between the input **x** and output 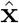. It quantifies how well the model captures underlying patterns. A low reconstruction error suggests the input conforms to learned normative distributions. High error indicates structural deviation [32] [33] and forms the basis of the sleep abnormality score:

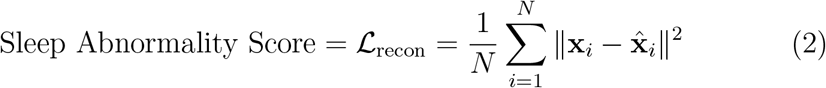

where ℒ_recon_ is the reconstruction loss, *N* is the number of features or time steps, **x**_*i*_ is the input feature vector, 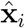 is the reconstructed vector, and ∥ *·* ∥^2^ is the squared Euclidean norm, which measures the direct geometric distance or structural deviation between the actual hypnogram and its reconstructed version.

The input to the autoencoder is a one-hot encoded hypnogram with dimensions (subjects, epochs, classes). Training data consisted of hypnograms from the combined SHHS and Sleep-EDF datasets, truncated or zero-padded to 1200 epochs (10 hours). Testing was performed on the WSC dataset, which includes multiple recordings per subject over a four-year span, enabling assessment of sleep score stability with age. Age is included to account for known changes in sleep patterns across the lifespan and to support longitudinal analysis. The model was trained using reconstruction loss.

#### 2.5.2. Model Architecture

Network architectures that directly model hypnogram data remain underexplored. Traditional time series models designed for continuous signals such as ECG or speech are poorly suited to hypnograms, which consist of discrete sleep stage transitions over long temporal sequences. We employed a Temporal Convolutional Autoencoder (TCN-AE) [34] to learn compressed representations of hypnograms by capturing both short- and long-range dependencies via dilated causal convolutions. Dilated convolutions allow the receptive field to expand exponentially with depth, enabling the model to learn temporal structures spanning the full night. The causal component ensures that temporal ordering is preserved, avoiding information leakage from future time steps. This design enables the encoder to extract multi-scale features and generate a latent space that retains both local and global sleep dynamics.

The autoencoder consists of four encoder blocks and four decoder blocks. In the encoder, each block contains a dilated convolution layer with dilation rate *q*_*l*_ = 4^*l*−1^, 64 filters with a kernel size of 8, Rectified Linear Unit (ReLU) activation, a channel reduction convolution to 16 channels, and average pooling for temporal downsampling. Skip connections are included to preserve information across layers. The subject’s age, normalised to the range [0, 1], is concatenated into the latent representation to account for age-related differences in sleep architecture. The decoder mirrors the encoder in reverse. Each block applies dilated convolutions with *q*_*l*_ = 4^4−*l*^, followed by ReLU activations and upsampling. A final convolutional layer restores the output to the original hypnogram dimensions. The full model architecture is illustrated in Figure 4.

**Figure 4:**
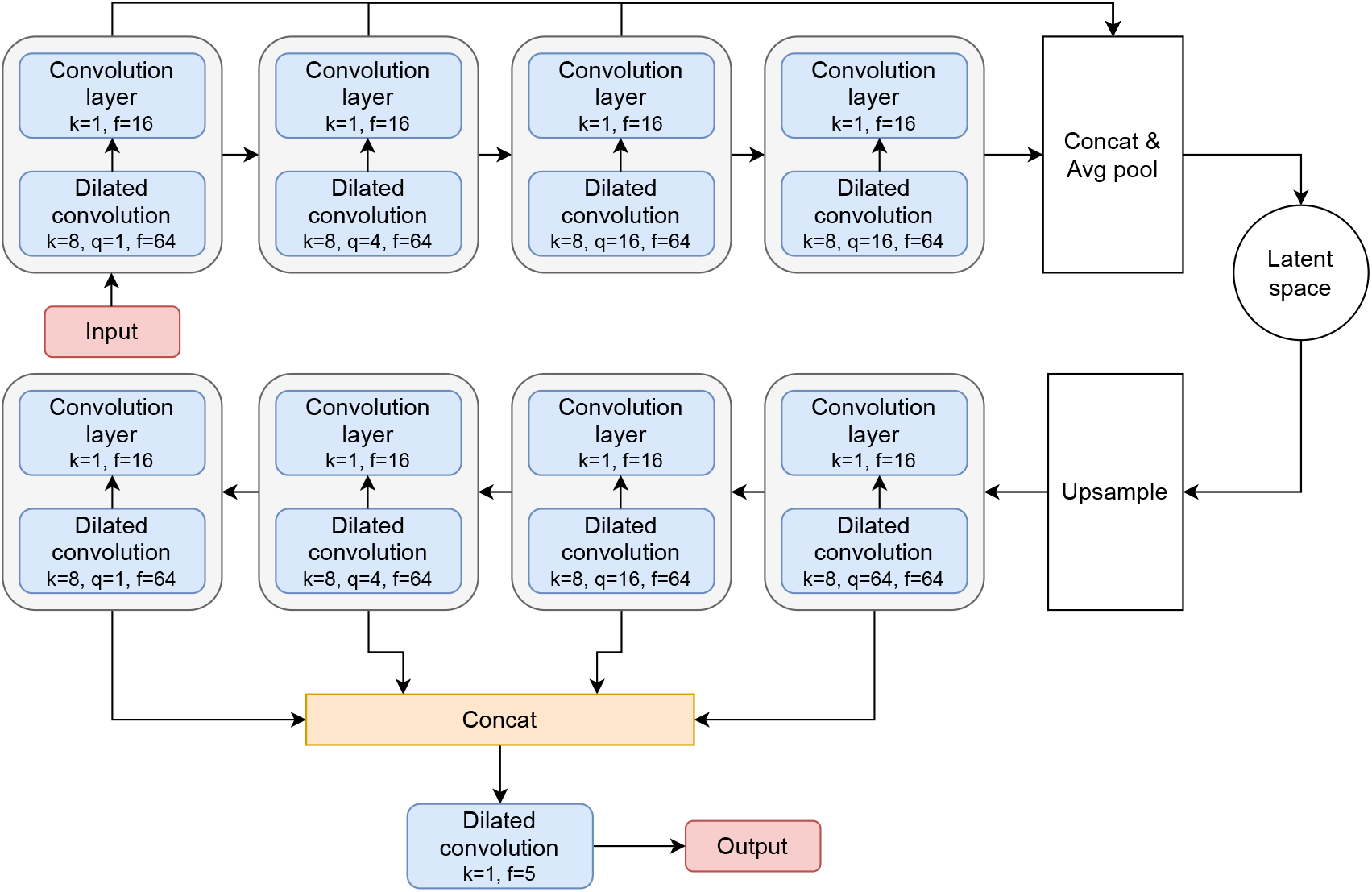
TCN-AE model for sleep abnormality scoring adapted from [34].

### 2.6. Performance Evaluation

#### 2.6.1. Sleep stage classification

We report metrics commonly used to evaluate sleep classification methods - accuracy (ACC) Eq. 3, F1 score (F1) Eq. 4 and the Cohen’s kappa (*κ*), which measures the agreement between the model and human raters [35].

To ensure fair comparison and isolate model performance, ensemble evaluation is based on single-channel training, with the same metrics reported.

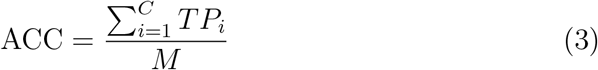

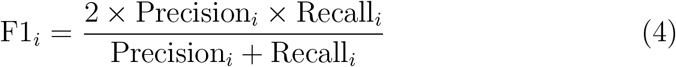

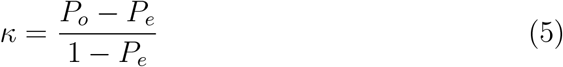

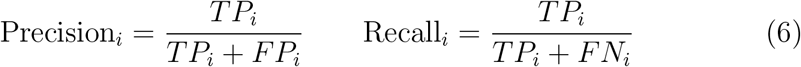

where *TP*_*i*_, *FP*_*i*_, and *FN*_*i*_ are the true positives, false positives, and false negatives for class *i* respectively, *C* is the total number of classes, and *M* is the total number of epochs, *P*_*o*_ is the observed agreement, and *P*_*e*_ is the hypothetical probability of chance agreement, calculated as:

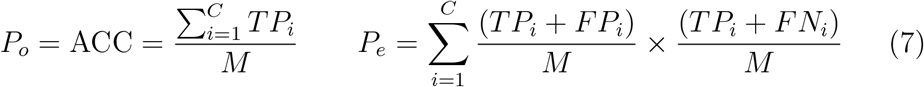

#### 2.6.2. Sleep scoring

Due to the lack of ground truth annotations for hypnograms, we evaluated the model-generated sleep abnormality score by correlating it with established clinical and behavioural sleep metrics. Our primary reference metric is the AHI [5], a widely used indicator of sleep-disordered breathing. A lower abnormality score is expected for subjects with normal sleep (AHI*<*5), and a higher score for those with abnormal or fragmented sleep. We computed the correlation between the model-derived abnormality score and the AHI values provided for each sleep recording.

In addition, we compared the abnormality score against components of the PSQI, a validated instrument for assessing subjective sleep quality. Higher PSQI scores indicate poorer sleep quality. We derived key PSQI-related metrics directly from the hypnogram:

##### Total Sleep Time (TST)

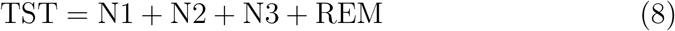

where N1, N2, and N3 are the durations of the three NREM sleep stages, and REM is the duration of the Rapid Eye Movement sleep stage.

##### Sleep Efficiency (SE)

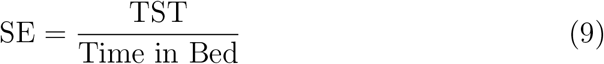

##### Sleep Disturbance (SD)

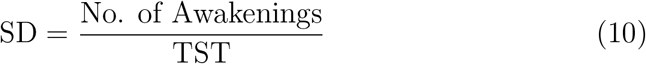

Spearman’s rank correlation coefficient *ρ* was used to evaluate associations between the abnormality score and sleep metrics.

Qualitatively, we assessed the model’s behaviour by introducing controlled distortions into valid hypnograms and observing changes in the abnormality score. Sample sleep recordings from the training set were selected, and their structure was progressively degraded by randomly altering the sleep stages of consecutive epochs in batches of ten. These modifications intentionally distort typical sleep progression patterns (e.g., transitions from N1 to REM), thereby disrupting the temporal integrity of the hypnogram.

Two percent of the training data was used for this evaluation. For each level of degradation, we computed the mean abnormality score across the selected subset. This procedure tests the sensitivity and monotonicity of the model’s output in response to increasingly abnormal sleep patterns. We expect the abnormality score to increase with greater distortion, demonstrating the model’s ability to detect deviations from physiologically realistic sleep architecture. An illustration of the progressive degradation procedure is shown in Figure 5.

**Figure 5:**
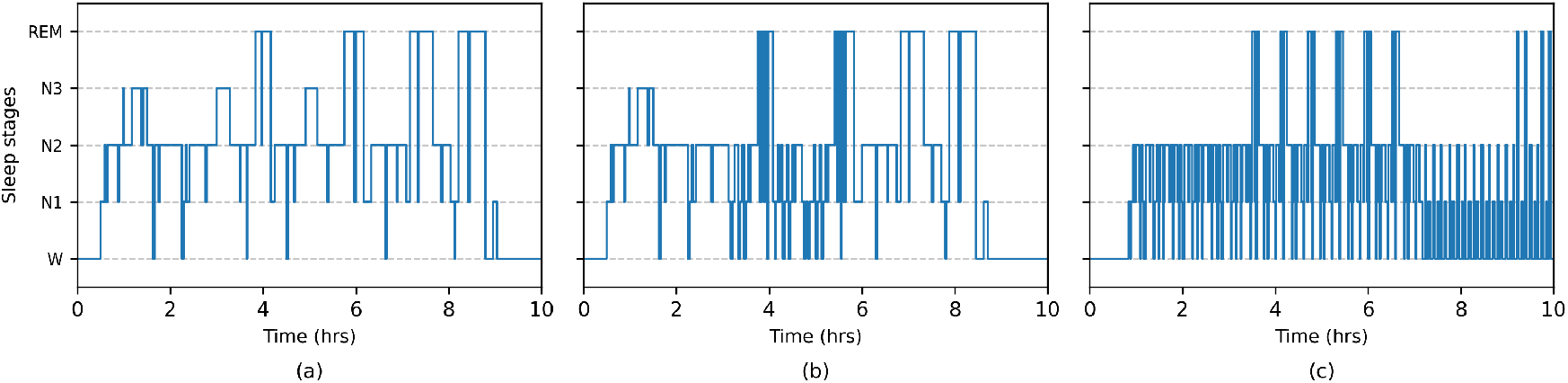
Illustration of a hypnogram with progressive degradation. (a) Original hypnogram. (b) Mildly fragmented hypnogram. (c) Highly fragmented hypnogram.

To complement the controlled degradation analysis, we examined representative real hypnograms from the held-out WSC test set. Two age-matched subjects were selected, and their hypnograms and standard sleep metrics (TST, SE, SD) were reported alongside the model-derived abnormality score.

## 3. Results

### 3.1. Sleep stage classification

#### 3.1.1. Base model results

Performance metrics for the deep learning architectures for multimodal tests are shown in Table 1. Given ChronoNet’s superior performance among non-attention architectures, we investigated whether injecting an attention mechanism could further boost its performance to match that of attention-based models. We implemented three modified versions of the ChronoNet model, where a CBAM layer is placed at the end of one of the three convolutional blocks - L1 (beginning), L2 (middle) or L3 (end). See Figure 6.

**Table 1:** Performance of deep learning architectures for multimodal tests.

| Model | ACC | F1 Score | | | | | $\kappa$ |
| --- | --- | --- | --- | --- | --- | --- | --- |
|  |  | W | N1 | N2 | N3 | REM |  |
| ChronoNet | 0.809 | 0.897 | 0.398 | 0.850 | 0.794 | 0.802 | 0.742 |
| + CBAM L1 | 0.755 | 0.834 | 0.316 | 0.783 | 0.771 | 0.733 | 0.672 |
| + CBAM L2 | 0.803 | 0.891 | 0.355 | 0.838 | 0.821 | 0.811 | 0.733 |
| + CBAM L3 | <u>0.839</u> | <u>0.907</u> | **0.466** | <u>0.891</u> | *0.848* | *0.817* | <u>0.767</u> |
| Hybrid-MultiChannelNet | 0.753 | 0.753 | 0.212 | 0.729 | 0.645 | 0.712 | 0.683 |
| SleepContextNet | *0.836* | *0.902* | 0.381 | *0.874* | <u>0.867</u> | **0.822** | 0.762 |
| ConvTransNet | 0.781 | 0.755 | 0.325 | 0.819 | 0.804 | 0.776 | 0.714 |
| **AttnSleep** | **0.848** | **0.911** | <u>0.452</u> | **0.894** | **0.881** | <u>0.820</u> | **0.784** |

**Figure 6:**
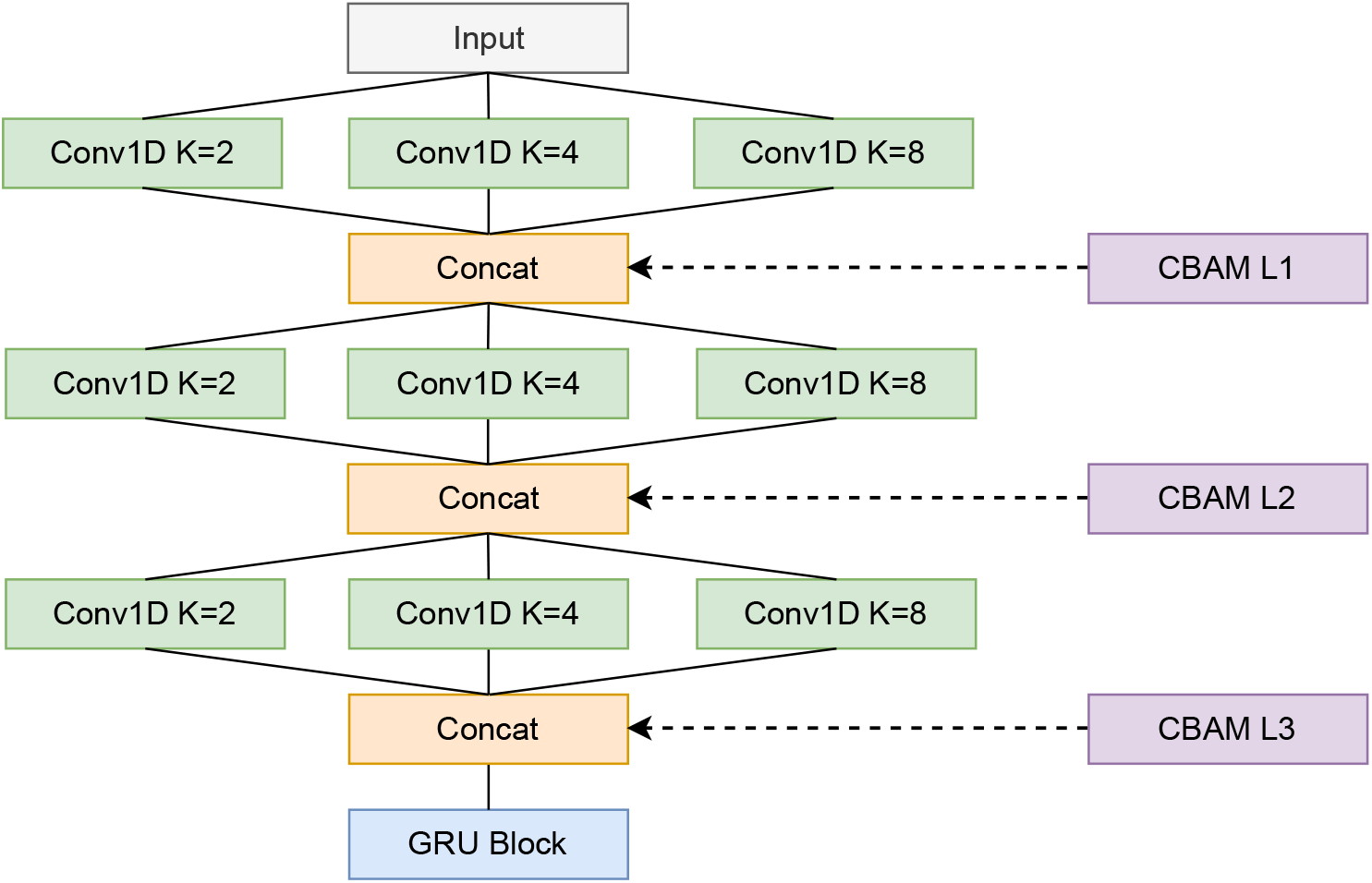
ChronoNet architecture [19] enhanced with CBAM [22] units for channel and spatial attention at the beginning, middle or final convolutional layers.

The results from the three highest-performing base architectures – ChronoNet, SleepContextNet, and AttnSleep – on single-channel EEG data are reported in Table 2.

**Table 2:** Single-channel test performance of the top three models from multimodal tests.

| Model | ACC | F1 Score | | | | | $\kappa$ |
| --- | --- | --- | --- | --- | --- | --- | --- |
|  |  | W | N1 | N2 | N3 | REM |  |
| ChronoNet | 0.721 | 0.781 | 0.302 | 0.743 | 0.757 | 0.729 | 0.672 |
| SleepContextNet | 0.800 | 0.874 | 0.336 | 0.814 | 0.735 | 0.761 | 0.741 |
| AttnSleep | 0.825 | 0.907 | 0.410 | 0.845 | 0.734 | 0.816 | 0.756 |

While absolute performance may vary across datasets due to differences in EEG channel placement and cohort characteristics, the aggregated results show consistent relative performance across models and between multimodal and single-channel settings, supporting the robustness of the reported findings.

#### 3.1.2. Ensemble model results

Based on the base model evaluations, AttnSleep and the attention-modified ChronoNet architectures demonstrated superior performance in sleep stage classification. As detailed in Section 2.4, we developed sleep stage-biased variants of these models to specialise in recognising a distinct sleep stage, forming the base learners for our ensemble framework. The performance metrics of these biased models, trained on single-channel data, are presented in Table 3. The performance of the ensemble models built on top of these stage-biased models is shown in Table 4.

**Table 3:** Performance metrics of the biased models for single channel (EEG) tests.

| Model | Class | ACC | F1 Score | | | | | $\kappa$ |
| --- | --- | --- | --- | --- | --- | --- | --- | --- |
|  |  |  | W | N1 | N2 | N3 | REM |  |
| ChronoNet+CBAM L3 | Wake | 0.763 | 0.921 | 0.284 | 0.783 | 0.711 | 0.682 | 0.943 |
| AttnSleep | N1 | 0.669 | 0.775 | 0.522 | 0.759 | 0.749 | 0.783 | 0.623 |
| AttnSleep | N2 | 0.779 | 0.826 | 0.311 | 0.887 | 0.773 | 0.753 | 0.905 |
| ChronoNet+CBAM L3 | N3 | 0.731 | 0.812 | 0.297 | 0.756 | 0.899 | 0.796 | 0.908 |
| AttnNet | REM | 0.756 | 0.813 | 0.320 | 0.772 | 0.767 | 0.841 | 0.840 |

**Table 4:** Performance metrics of the ensemble models for single channel (EEG) tests.

| Model | ACC | F1 Score | | | | | $\kappa$ |
| --- | --- | --- | --- | --- | --- | --- | --- |
|  |  | W | N1 | N2 | N3 | REM |  |
| Encoder-only ensemble | 0.796 | 0.909 | 0.397 | 0.832 | 0.809 | 0.716 | 0.721 |
| Encoder-Decoder ensemble | 0.805 | 0.894 | 0.444 | 0.825 | 0.814 | 0.819 | 0.753 |

### 3.2. Sleep Scoring

For subjects in the training and test datasets, the range of abnormality scores was between [1.01, 1.21], with the mean being 1.09. Across the WSC test set, predicted hypnograms from the best-performing classification model showed consistently higher scores, with an average increase of 0.154 (SD: 0.0373) compared to their ground-truth counterparts. Figure 7 shows the plot of the abnormality scores against each of the individual metrics from the PSQI. Spearman’s coefficient for the correlation between the abnormality score and sleep efficiency is -0.436 with a *p*-value of 0.00024. Spearman’s coefficient for the correlation between SD and abnormality score is 0.33 with a *p*-value of 0.0014.

**Figure 7:**
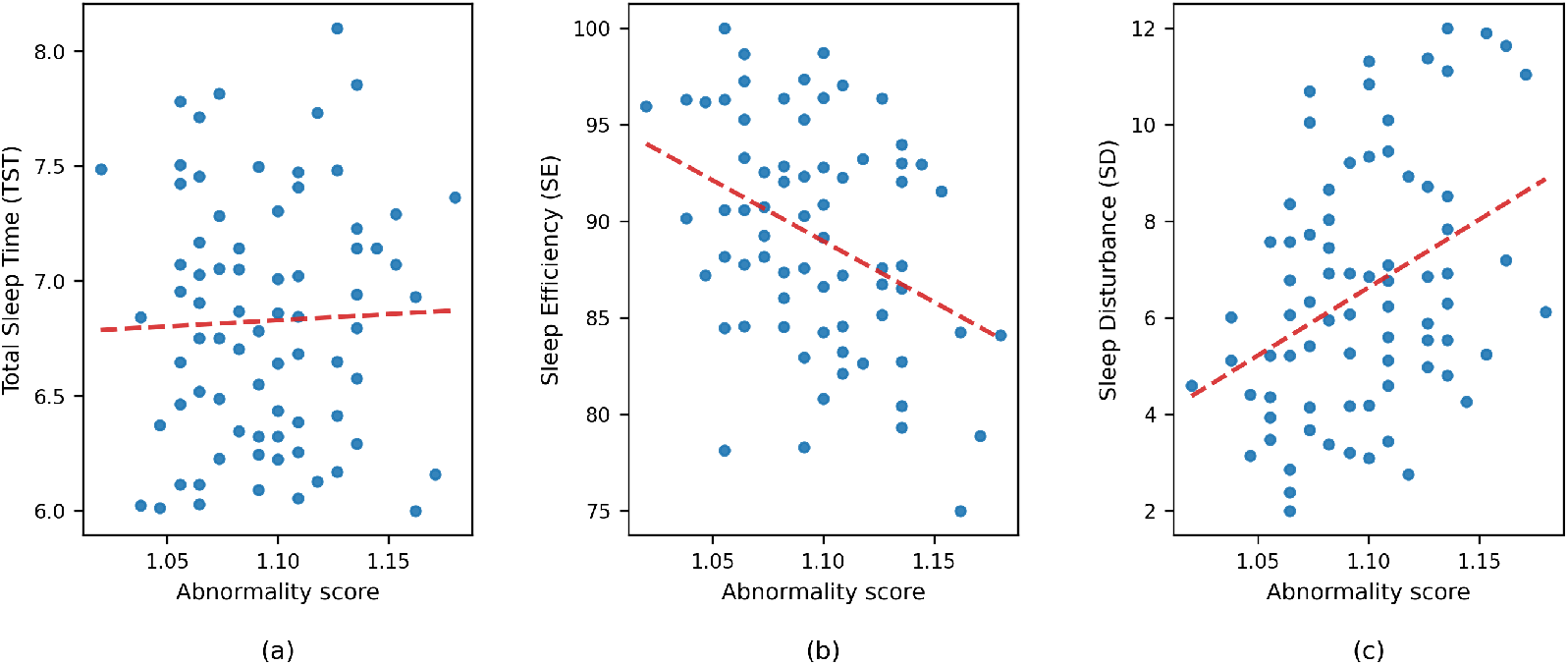
Sleep abnormality score vs PSQI metrics. (a) Abnormality score vs Total Sleep Time (TST). (b) Abnormality score vs Sleep Efficiency (SE). (c) Abnormality score vs Sleep Disturbance (SD).

Figure 8 shows the abnormality score vs AHI and age scatter graphs for the model. Figure 9 depicts the change in the abnormality score by introducing controlled degradation to the hypnogram.

**Figure 8:**
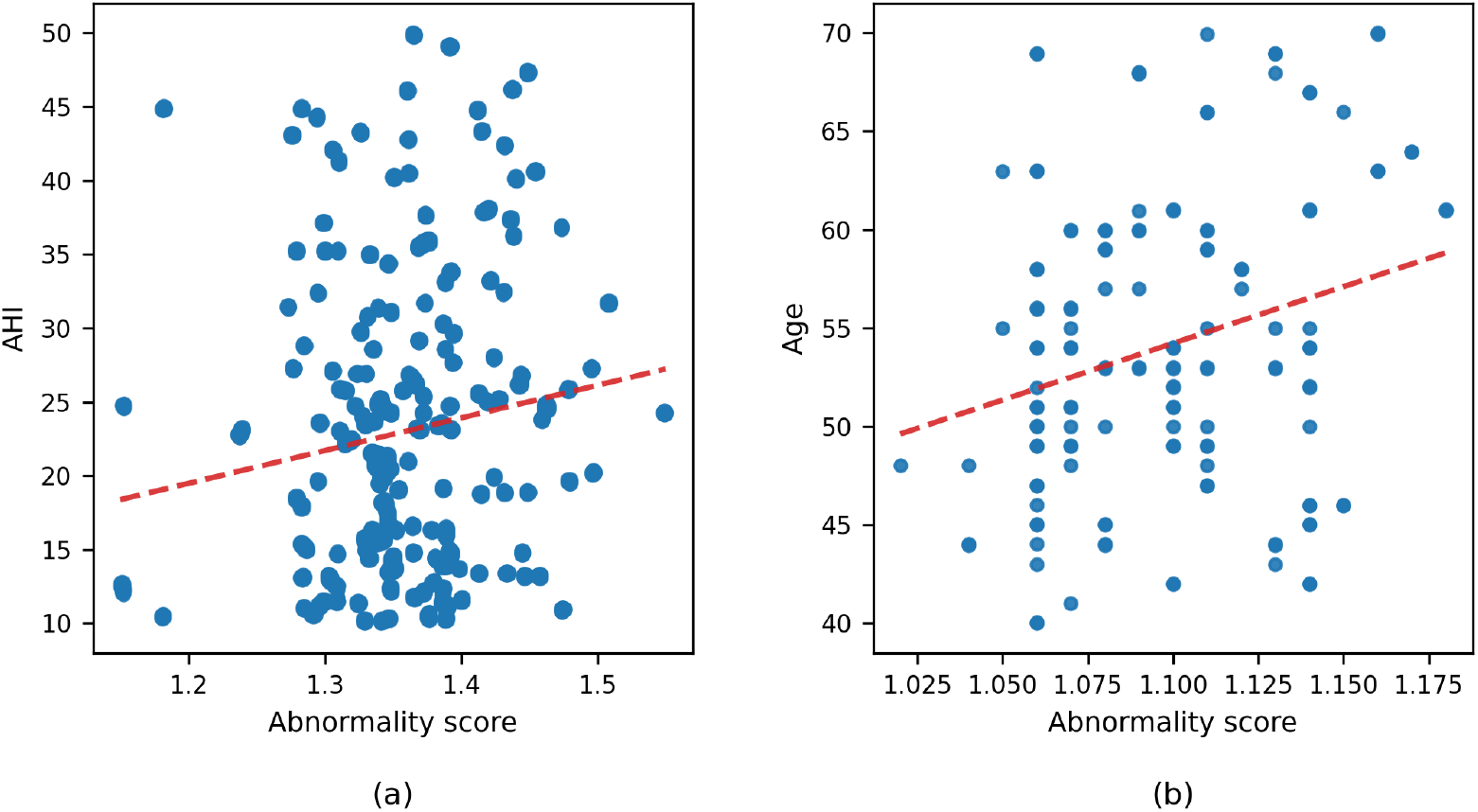
Sleep abnormality score vs (a) Apnoea-Hypopnoea Index (AHI), and (b) Age.

**Figure 9:**
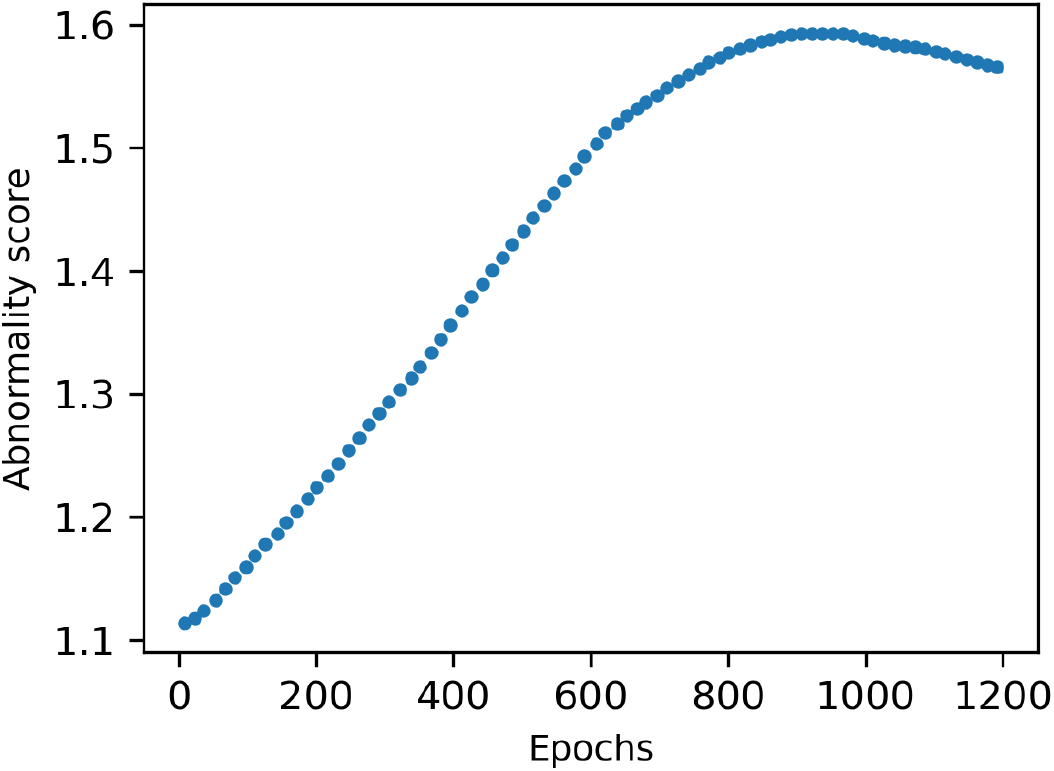
Evolution of sleep abnormality score with controlled hypnogram degradation.

To illustrate the behaviour of the abnormality score on real-world data, Table 5 outlines the sleep metrics for two representative cases from the WSC test set, consisting of two age-matched subjects between 45-50 years with AHI values of 11.43 and 44.18. Figure 10 presents the corresponding ground-truth hypnograms.

**Table 5:**
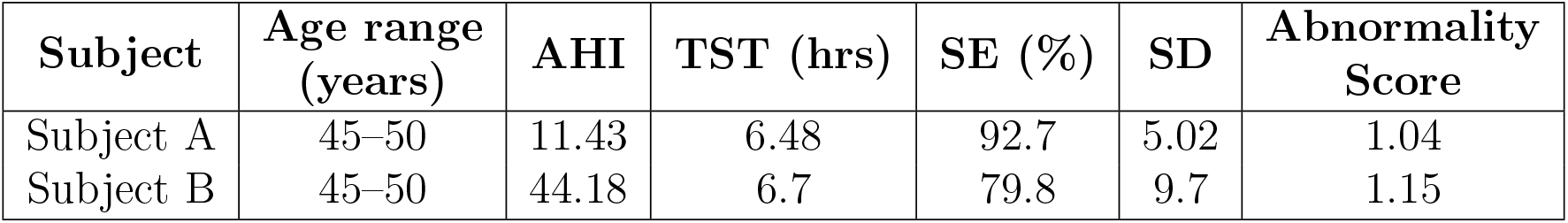
Representative age-matched WSC test cases illustrating the relationship between clinical sleep metrics and the proposed abnormality score.

| Subject | Age range (years) | AHI | TST (hrs) | SE (%) | SD | Abnormality Score |
| --- | --- | --- | --- | --- | --- | --- |
| Subject A | 45–50 | 11.43 | 6.48 | 92.7 | 5.02 | 1.04 |
| Subject B | 45–50 | 44.18 | 6.7 | 79.8 | 9.7 | 1.15 |

**Figure 10:**
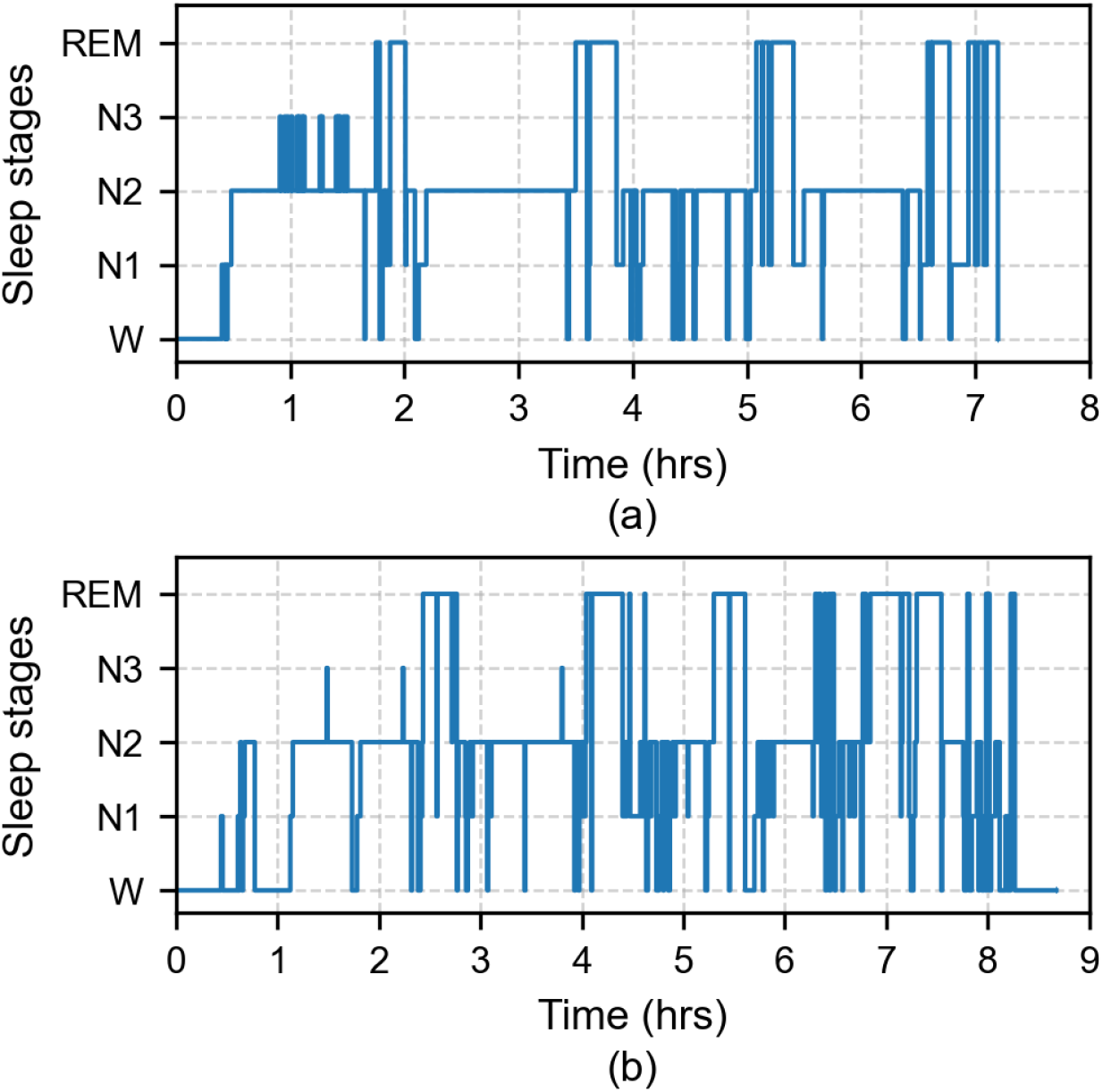
Representative hypnograms from age-matched WSC test subjects illustrating differences in sleep architecture associated with abnormality severity. Subject A exhibited mild sleep-disordered breathing (AHI = 11.43), preserved sleep efficiency (92.7%), and a lower abnormality score (1.04), with relatively stable transitions between sleep stages. In contrast, Subject B demonstrated severe sleep-disordered breathing (AHI = 44.18), reduced sleep efficiency (79.8%), increased sleep disruption (SD = 9.7), and a higher abnormality score (1.15), reflected by frequent awakenings and fragmented sleep-stage transitions.

## 4. Discussion

Attention-based models for sleep stage classification, particularly AttnSleep and attention-enhanced ChronoNet, demonstrated strong performance in classifying sleep stages using multimodal physiological signals (EEG, EMG, and EOG). Nevertheless, classification accuracy remains affected by significant class imbalance and the inherent difficulty in distinguishing between physiologically similar stages, especially N1. This is due to its transitional nature between wakefulness and deeper sleep, making it highly susceptible to misclassification with either Wake or N2. As expected, all models exhibited performance degradation when fewer input channels were available, highlighting the challenge of learning discriminative features from reduced data. ChronoNet, which was specifically designed for EEG data, experienced a notable performance drop under these constraints. This could stem from its specialised EEG processing being optimised for tasks where labels are implicitly informed or validated by multimodal data. This warrants further investigation. In contrast, attention-based models were more robust to the removal of input channels, showing smaller performance losses than their non-attention-based counterparts.

The addition of attention mechanisms such as CBAM improved ChronoNet’s performance. The placement of attention within the network is important. Applying CBAM at the input stage yields negligible learning and degrades overall performance, likely because attention is applied before the convolutional layers have extracted meaningful temporal features. When CBAM is positioned mid-network, the effect is minimal. However, inserting CBAM after the final convolutional block leads to a noticeable improvement in classification performance. This strategic late-stage placement allows the module to selectively re-weight the most discriminative temporal features already extracted by the network, just before the subsequent recurrent or fully connected layers make the final classification decision. The modified ChronoNet with CBAM placed at the end achieved the highest F1-score for N1 classification across all models tested in both multimodal and single-channel configurations. Despite this improvement however, the F1-score for N1 remained below 0.5, underscoring the persistent challenge of accurately detecting this ambiguous stage. This is consistent with prior work, as N1 represents a transitional stage with weak and heterogeneous electrophysiological signatures, often overlapping with both wakefulness and N2, making it inherently difficult to classify [36] [37].

The ensemble models were developed to enhance classification performance across sleep stages, motivated by the observation that single models often struggle to learn discriminative spatiotemporal patterns due to heterogeneous physiological signatures. The proposed strategy leverages multiple base models, each biased toward a specific sleep stage, which are then combined via a stacking-based ensemble to correct misclassifications and integrate complementary strengths. Each biased model showed improved performance on its respective target class compared to non-biased counterparts. For example, the N1-specific biased model achieved a class-specific F1-score slightly above 0.5, markedly better than general-purpose models, despite N1 remaining the most challenging class to classify accurately.

As shown in Table 4, both the encoder-only and encoder-decoder ensemble models outperformed the individual stage-biased base models from which they were constructed, in terms of overall classification accuracy, confirming the benefit of aggregating specialised classifiers. However, compared to strong standalone architectures such as AttnSleep, the ensemble achieves comparable but not superior overall performance. The encoder-decoder ensemble achieved the highest accuracy of 0.805, exceeding the encoder-only variant, while individual biased model accuracies ranged from 0.669 to 0.779. These results support the intuition that aggregating specialised models improves generalisability and mitigates individual model biases. The primary advantage of the ensemble lies in improved class-specific performance, particularly for the challenging N1 stage (0.444 vs 0.410 in AttnSleep, Table 2), rather than in overall accuracy gains.

The ensemble-driven gains are not uniformly distributed across sleep stages. While the encoder-decoder ensemble achieved the highest F1-score for N1 (0.444), its performance on N2, N3, and REM showed only marginal differences. For instance, although it attained strong F1-scores for N2 (0.825) and REM (0.819), these are comparable to those of AttnSleep (0.845 for N2 and 0.816 for REM). This may be because the representational power of the ensemble remains bounded by the quality and diversity of the base models it aggregates. These results suggest that ensemble learning is particularly effective for challenging stages such as N1, where specialised models provide complementary decision boundaries, but offers limited gains for well-separated stages.

A positive correlation was observed between sleep duration and the abnormality score, while a negative correlation emerged between sleep efficiency and the abnormality score, both consistent with expectations. No significant correlation was found between the abnormality score and total sleep time. Note that recordings exceeding 10 hours were excluded and very few test subjects exhibited substantially short sleep durations.

According to SHHS and WSC guidelines, an AHI *<* 5 is considered indicative of normal sleep, with higher values reflecting increasing sleep pathology. While elevated AHI is associated with disrupted sleep architecture, including increased fragmentation and altered stage distribution [5] [38] [39], the relationship is indirect and heterogeneous across individuals. In our results, no strong monotonic relationship was observed between AHI and the abnormality score (Figure 8). This may reflect the fact that AHI quantifies the frequency of apnoea and hypopnoea events, whereas the proposed abnormality score captures deviations in the temporal structure of sleep stages. Although these phenomena are related, they do not always align directly, particularly in cases where respiratory events do not substantially alter global hypnogram structure. This suggests that the proposed score captures complementary aspects of sleep disruption not fully reflected by AHI alone.

As discussed in Section 2.5, it is essential that the abnormality score remains independent of age. Figure 8 shows limited correlation between the two. Although Spearman’s coefficient is 0.156, the p-value of 0.0946 suggests this correlation is not statistically significant, supporting the model’s intended age independence. While age is incorporated into the latent space to account for population-level variability, we do not perform an explicit ablation study. Evaluating the isolated contribution of age conditioning is left for future work.

When hypnograms are progressively degraded through controlled alterations, the model produces steadily increasing abnormality scores. An initially linear relationship between degradation and score is observed, which eventually plateaus before full hypnogram degradation (see Figure 9). Beyond this point, a slight decline in the score is noted, suggesting a saturation effect. This behaviour indicates the model’s sensitivity to structural sleep disruptions and its ability to detect abnormality in a graded manner. This is consistent with prior work showing that sleep fragmentation and disrupted stage transitions are key indicators of abnormal sleep architecture [38, 39]. Further, our illustrative cases from the WSC cohort show that higher AHI and increased sleep disruption (lower SE and higher SD) are consistent with higher abnormality scores.

Methods designed specifically for modelling hypnograms are still limited, as most conventional time-series architectures are built for continuous signals rather than the discrete, stage-based transitions that characterise sleep. Our results demonstrate that the proposed sleep abnormality score provides an objective, interpretable measure of deviations from typical sleep architecture. Higher scores indicate greater abnormality, and the metric’s weak dependence on age makes it particularly suitable for longitudinal monitoring in at-risk populations. The score’s sensitivity to artificially degraded hypnograms further confirms its ability to capture subtle disruptions in sleep structure. Although scores computed from model-predicted hypnograms are systematically higher than those from ground truth, this upward shift likely reflects structural differences encoded by the autoencoder. Importantly, the relative variations among predicted scores remain meaningful, supporting their use as a label-free proxy for sleep disruption within end-to-end automated pipelines. This aligns with prior studies emphasising the importance of objective, structure-based measures for assessing sleep quality beyond subjective or event-based metrics [5].

While the proposed framework demonstrates promising results, several limitations should be considered. First, the abnormality score is not validated against disease-specific clinical diagnoses and should be interpreted as a structure-based indicator of sleep disruption rather than a diagnostic measure. Second, validation relies on proxy metrics such as PSQI components and AHI, as well as controlled hypnogram degradation, which do not fully capture the complexity of real-world sleep pathology. Third, variability in EEG channel configurations and cohort characteristics across datasets may influence performance. In particular, the use of different EEG derivations across studies limits direct comparability and may introduce variability that would be reduced under a standardised recording montage. Finally, although age is incorporated into the latent representation, an explicit ablation study to isolate its contribution was not performed and remains an area for future investigation.

In the context of existing literature, most prior approaches focus on supervised sleep stage classification or subjective sleep quality assessment. The proposed method provides a complementary, label-free measure of sleep structure based on hypnogram dynamics. Its clinical utility remains to be established through validation on clinically annotated cohorts and disorder-specific populations.

## 5. Conclusion

This study presents a framework for label-free quantification of sleep abnormality from hypnograms, supported by deep learning-based sleep stage classification and ensemble modelling. The proposed approach enables objective assessment of sleep structure directly from hypnograms using large-scale public datasets. Collectively, these contributions represent foundational steps toward a multimodal, end-to-end sleep analysis framework, offering a scalable, low-resource alternative to conventional vPSG-based monitoring. Unlike traditional tools such as the PSQI, which are prone to subjectivity and recall bias, the proposed framework enables continuous, objective, and long-term sleep assessment, with potential relevance for monitoring sleep-related changes in clinical populations.

Future work should investigate the integration of the abnormality scoring mechanism into classification pipelines to enable continuous, adaptive monitoring. Accounting for contextual variables such as circadian rhythms, ambient light, and environmental noise could further enhance model accuracy and relevance. Additionally, adapting the framework for wearable-device data through denoising and signal quality enhancement strategies will be essential for scalable deployment in real-world, home-based settings. Finally, the development of lightweight, interpretable models will improve the accessibility and transparency of automated sleep analysis, supporting broader adoption in clinical and remote health applications.

## Data Availability

The data used in this study were obtained from the National Sleep Research Resource (NSRR). Access is available to qualified researchers upon application to and approval by NSRR, subject to a data use agreement. The authors are not permitted to redistribute the data.

https://sleepdata.org/datasets/shhs

https://sleepdata.org/datasets/wsc

https://physionet.org/content/sleep-edfx/1.0.0/

## 6. CRediT authorship contribution statement

**Zekeriye Nur**: Conceptualization, Data curation, Methodology, Software, Visualization, Writing - Original Draft. **Nivedita Bijlani**: Visualization, Writing - Review & Editing. **Mauricio Villarroel**: Conceptualization, Validation, Resources, Supervision, Funding Acquisition.

## 9. Data availability

The data used in this study are available from the National Sleep Research Resource (NSRR) and PhysioNet repositories. Specifically, we used the following datasets:

1. Sleep Heart Health Study (SHHS): https://sleepdata.org/datasets/shhs
2. Wisconsin Sleep Cohort (WSC): https://sleepdata.org/datasets/wsc
3. Sleep-EDF-20 and Sleep-EDF-78: https://physionet.org/content/sleep-edfx/1.0.0/

Access to these datasets may require user registration and approval depending on the dataset-specific data use agreements.

